# Do Must-Access Prescription Drug Monitoring Programs (PDMPs) Affect Pain and Physical Impairment Outcomes in Older Adults?

**DOI:** 10.1101/2025.06.02.25328258

**Authors:** Martha Wetzel, David H Howard, Nicholas A Giordano, Stephen W Patrick, Courtney R Yarbrough

## Abstract

**Background:** State policies requiring clinicians to review prescription drug monitoring program (PDMP) databases have proliferated. Patient advocates warn that such policies may reduce quality of life for some patients.

**Objectives:** To quantify the relationship between must-access PDMPs and pain and physical impairment outcomes.

**Research Design:** Using Health and Retirement Survey (HRS) data on older adults in the US from 2002-2021, we conducted a heterogeneity-robust difference-in-differences analysis.

**Results:** The population included 34,431 individuals with an average age of 67. Must-access PDMPs were initially associated with 1.65 (95% CI: 0.43 - 2.87) to 3.52 (95% CI: 0.88 - 6.16) percentage point increases in the reports of frequent pain. However, the effect dissipated over time. Effects on the impairment outcomes were positive but statistically insignificant.

**Conclusions:** Must-access PDMP policies are associated with an initial increase in frequent pain reported by older adults. No significant effects on physical impairment outcomes were found. Continued research examining the role of PDMPs on patient health outcomes is warranted.

## Introduction

The opioid crisis and the high prevalence of pain in the US are fundamentally linked, with physical pain contributing to opioid use and dependence (Fornili, 2018). Although the National Academies of Science recommends a comprehensive public health response addressing both pain and opioids, popular and political attention has focused on restricting the opioid supply (Goodin, 2018; Gross & Gordon, 2019; Holmgren et al., 2020; National Academy of Sciences, 2017). One of the most common state-level responses has been the implementation of prescription drug monitoring programs (PDMPs). PDMPs are statewide databases documenting filled prescriptions for controlled substances, which can then be checked by both prescribers and pharmacists. Evaluations of these programs have focused primarily on the intended, supply-related outcomes, such as opioid prescribing rates (Tay et al., 2023). However, patient-centered outcomes, such as quality of life or functional status, are largely absent from the PDMP literature (Hoppe et al., 2022; Tay et al., 2023) as well as the broader opioid policy literature (Coleman et al., 2023). One literature review identified over 150 studies on PDMPs and concluded that, “Research on individual patient health outcomes, for example…pain and mental health status are notably lacking.” (Hoppe et al., 2022) In this study, we aim to fill this knowledge gap by assessing the relationship between must-access PDMPs and pain- and impairment-related outcomes in older adults. These results will provide a deeper understanding of functional outcomes associated with PDMPs, which can inform modifications to existing PDMPs and targeted clinical interventions when PDMPs go into effect.

Both opioid misuse and pain are costly, prevalent issues. The Centers for Disease Control and Prevention (CDC) estimates that approximately 50 million adults in the US suffer from chronic pain, with 19.6 million of them experiencing high-impact chronic pain that limits life or work activities most days (Dahlhamer et al., 2018). Among older adults, pain is particularly common, with 28 percent of men and 35 percent of women ages 55 and up reporting frequent pain (Zimmer & Zajacova, 2020). One consequence of pain is decreased functional capacity. Adults in their 50s who have pain report similar levels of functioning to adults in their 80s who do not report pain (Covinsky et al., 2009). Consequently, the economic burden of chronic pain is estimated to be $560 billion, higher than heart disease and stroke (Benjamin et al., 2018; Gaskin & Richard, 2012). Opioid use disorder is also common and costly, affecting 5.7 million people in the US over age 12 (Substance Abuse and Mental Health Services Administration, 2024), with an estimated economic impact over $1 trillion annually (Florence et al., 2021).

One factor that contributed to the rise in opioid use disorders was the movement to treat pain aggressively with opioids in the 1990s and 2000s (Tompkins et al., 2017). Following a rapid expansion in opioid prescribing and opioid overdose deaths (Tompkins et al., 2017), the CDC released guidelines that recommended nonopioid over opioid treatment for chronic pain in 2016 (Dowell, Haegerich, et al., 2016). The guidelines highlighted the unclear benefits and known risks of long-term opioid use (Dowell, Haegerich, et al., 2016). For conditions such as chronic low back pain, clinical trials have shown that opioids do not outperform NSAIDS in terms of pain control (White et al., 2011). Based on this literature, one could reasonably expect reductions in opioid prescribing at the population level to translate to neutral or improved patient outcomes. However, since pain patients included in clinical trials are generally not representative of the average pain patient (Sehgal et al., 2013), real world results may differ from expectations. Additionally, this expectation assumes that providers will deliver alternative therapies to opioids and that patients have ready access to them, which may not prove true.

Substantial research has explored changes to clinical practice, including possible harms from discontinuation of opioid therapy for patients in chronic pain, following the 2016 CDC guidelines (Darnall et al., 2019; Dowell et al., 2022; Fenton et al., 2019; Peachman, 2023). In contrast, limited literature explores changes to clinical practice and patient outcomes following PDMP implementation. The informal literature contains a narrative that decreasing opioid prescribing has harmed patients with chronic pain (Anson, 2017; Gleason, 2014; Human Rights Watch, 2018) and contributed to suicides among pain patients (Szalavitz, 2022). One qualitative study conducted focus groups with patients who took opioids following several opioid-related policy changes in Ontario, Canada, including formulary changes and a PDMP. Pain patients reported increased stigmatization, deteriorating relationships with providers, and increased difficulties in handling their health and pain (Antoniou et al., 2019). However, one quantitative study on opioid tapering following PDMPs yielded mixed results (Bao et al., 2021).

### Conceptual Model

We conceptualize PDMPs as laws that may affect multiple aspects of the health care environment using the public health law framework developed by Komro et al (Komro et al., 2013). Prior PDMP literature has proposed a conceptual model that considers changes in opioid prescribing as the driver of outcomes (Finley et al., 2017). We build on and expand that model to allow PDMPs to influence the pain care outcomes not only via the opioid prescriptions but also through changes to the clinician-patient interactions.

Clinicians consistently self-report reducing opioid prescribing because of PDMPs (Picco et al., 2021). In addition, the literature documents a variety of other clinician-reported practice changes in response to PDMPs, including changes in non-controlled substance prescribing, communication with patients and other providers, and stigma (Picco et al., 2021). For example, clinicians who reduce their opioid prescribing due to the PDMP may adapt by offering a reduced set of pain management options that excludes opioids or by omitting pain control discussion altogether.

Patient expectations and conditioning are known to influence pain treatment outcomes (Colloca, 2019). One opioid-stewardship training program found that clinicians who had undergone the training had lower patient trust scores compared to control clinicians (Sherman et al., 2018). For chronic pain patients, changes induced by the PDMP are occurring in a clinical context that may already be characterized by mutual clinician-patient frustration and distrust (Dobscha et al., 2008; Upshur et al., 2010). Thus, our conceptual model posits that PDMPs influence pain outcomes directly through opioid prescribing and indirectly through changes to the pain care experience.

### New Contribution

Although multiple commentaries have drawn attention to the importance of evaluating the effect of PDMPs on pain-related outcomes (Goodin, 2018; Gross & Gordon, 2019), PDMP research has primarily focused on opioid supply (Hoppe et al., 2022). The few papers that incorporate a pain-related outcome (Kilby, 2015; Wetzel et al., 2021) are focused on acute pain patients and rely on older difference-in-difference methods that are not robust to heterogenous effects. To our knowledge, this study is the first to employ heterogeneity-robust analytic methods to evaluate the association between must-access PDMPs and pain and physical impairment outcomes.

## Methodology

### Data Sources

In this retrospective cohort study, we used data from the Health and Retirement Study (HRS) from 2002 to 2020 (Health and Retirement Study, 2025). The HRS is an ongoing longitudinal survey of more than 37,000 noninstitutionalized adults ages 50 and above in the US (Health and Retirement Study, 2017; Sonnega et al., 2014). New cohorts are added every six years and follow-up surveys are conducted with participants every two years (Sonnega et al., 2014). The multi-stage area probability sample design, recruitment, follow-up, and data collection practices have been described elsewhere (Heeringa & Connor, 1995; Sonnega et al., 2014). The HRS is one of the longest-running studies that includes a suite of questions focusing on disability, functioning, and pain (Agree & Wolf, 2017).

The independent variable of interest is the presence of a prescriber must-access PDMP law. We define must-access PDMP laws as those that require the prescriber to check the PDMP prior to issuing an initial opioid and at regular intervals thereafter. We do not consider laws that allow exceptions for clinician judgement, established patients, or prescriptions under 30 days supply to be must-access laws. In states that experienced implementation delays (e.g., Pennsylvania), we use the date that the system was functional rather than the date specified in the law. To develop the data set of PDMP laws, the authors cross-referenced existing PDMP law data sets (Horwitz et al., 2021; RAND-USC Schaeffer Opioid Policy Tools and Information Center, 2021b) with a compilation of mandatory use conditions published by the PDMP Training and Technical Assistance Center (Prescription Drug Monitoring Program Training and Technical Assistance Center, 2024). Then, the legal codes were referenced to verify implementation dates. The policy data was linked to the HRS survey data based on state; because observations missing state were excluded, the match was exact. During the time period 2002-2021, 43 states plus Washington, DC, implemented a must-access PDMP law (Supplemental Table 1).

Given that the HRS is conducted in two-year cycles, we count time relative to policy implementation in terms of survey waves. All interviews and policies were assigned to a survey wave, and interviews in treated states taking place during the same wave as policy implementation were assigned *t=0*.

This study was deemed exempt by the Emory University Institutional Review Board. The authors had access to the restricted data files containing state-level geographic identifiers.

### Outcomes

The outcomes included one measure of pain frequency and three measures of physical impairment. The frequent pain outcome was determined by a positive response to the question, “are you often bothered by pain?” The first measure of impairment, activity-limiting pain, was based on yes responses to the question “Does the pain make it difficult for you to do your usual activities such as household chores or work?” Two additional impairment measures were a count of bedridden days and a count of functional limitations. Bedridden days were defined as days spent in bed due to illness or injury in the past month. The count of functional limitations was based on responses to twelve questions asking about difficulty with physical functioning tasks related to mobility, strength, and motor skills, such as ability to jog a mile and ability to climb a set of stairs (Supplemental Table 2). Functional limitations and bedridden days were treated as continuous variables. A complete list of codes and data cleaning steps is provided in Supplemental Table 3. Observations missing an outcome made up less than 0.5 percent of the sample and were dropped on a model-by-model basis.

Descriptive measures include age, race, ethnicity, education, and insurance status. In 2016, a question about opioid use in the past three months was added to the HRS. Although there were not enough pre-policy data to include opioid use as a study outcome, we provide descriptive statistics on this measure for context. In addition, we report whether the observation is from a respondent living in a state with a medical marijuana law based on data published by the RAND-USC Schaeffer Opioid Policy Tools and Information Center (RAND-USC Schaeffer Opioid Policy Tools and Information Center, 2021a).

### Sample

We included all observations from participants that 1) resided in a state that implemented a must-access PDMP during the time period of interest, and 2) included a response to the frequent pain question. Observations missing state of residence or residing in a US territory (N= 707) were excluded. Responses from participants in late-treated states were used as the comparison group. The late-treated states were chosen as the comparison group over the never-treated states because the never-treated group was limited to seven states, six of which were located in the Midwest. As a sensitivity analysis, we reran the models using never-treated states as the comparison group instead of the late-treated states.

### Empirical Approach

To calculate average program effects, we employed a heterogeneity-robust difference-in-difference model with staggered treatment timing. We prepared the data by classifying the PDMP states into treatment cohorts based on the survey wave during which the PDMP was first implemented. We then use an outcome regression approach outlined by Callaway and Sant’Anna (2021), which subsets the data based on treatment cohort and time, estimates each cohort-time-specific treatment effect separately, and then aggregates the estimates. Each subset includes one treatment cohort and the later-treated controls at the time point of interest (τ) and the time point prior to the cohort’s PDMP implementation (*g-1*). The models had the following specification:

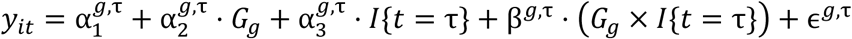

Where *G*_*g*_ indicates PDMP cohort was first treated at survey wave g and *I*{*t* = *τ*} is a post-treatment timing indicator. The binary outcomes of frequent pain and disabling pain were estimated using linear probability models. Standard errors were clustered at the state level. Simultaneous confidence bands were estimated via multiplier bootstrap. The average treatment effects on the treated (ATT) were aggregated to produce an overall ATT and dynamic effects that varied by time relative to PDMP implementation. These dynamic ATT estimates and accompanying 95% confidence intervals were used to assess the identifying assumption that the treated and untreated units had similar trends prior to treatment.

## Results

After excluding observations missing state of residence (N=707) and those missing a response to the frequent pain question (N=510), 187,881 observations representing 34,431 individuals remained. Post-treatment observations numbered 28,043 (14.9%). The mean age was 67 (SD: 11.4) and nearly 60 percent of the observations were from women (Table 1). Half of the states had both medical marijuana and must-access PDMP policies. Of these states with both policies, all but two implemented medical marijuana before the must-access PDMP, with an average of 9 years elapsing between medical marijuana and must-access PDMP implementation.

**Table 1:**
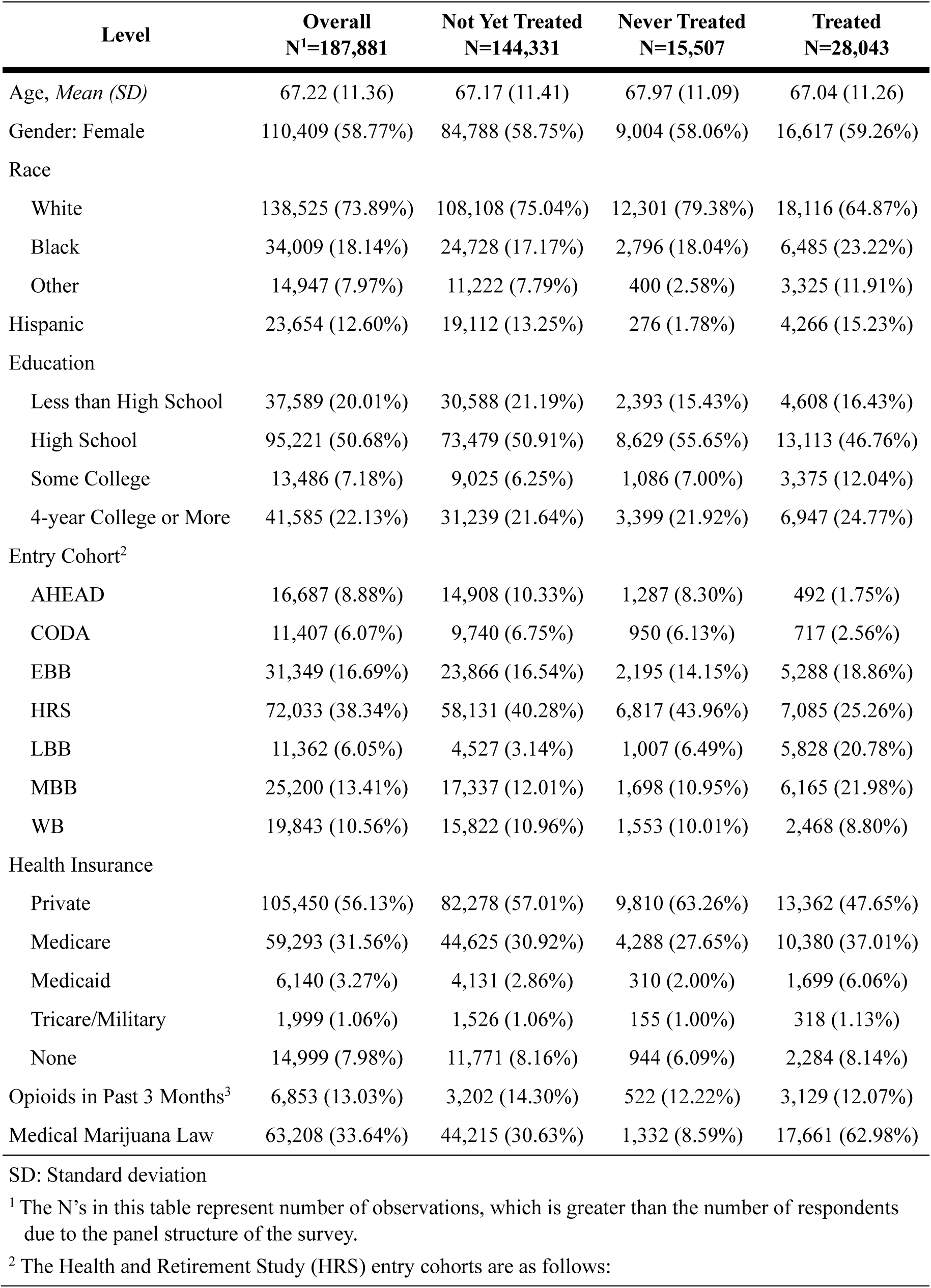
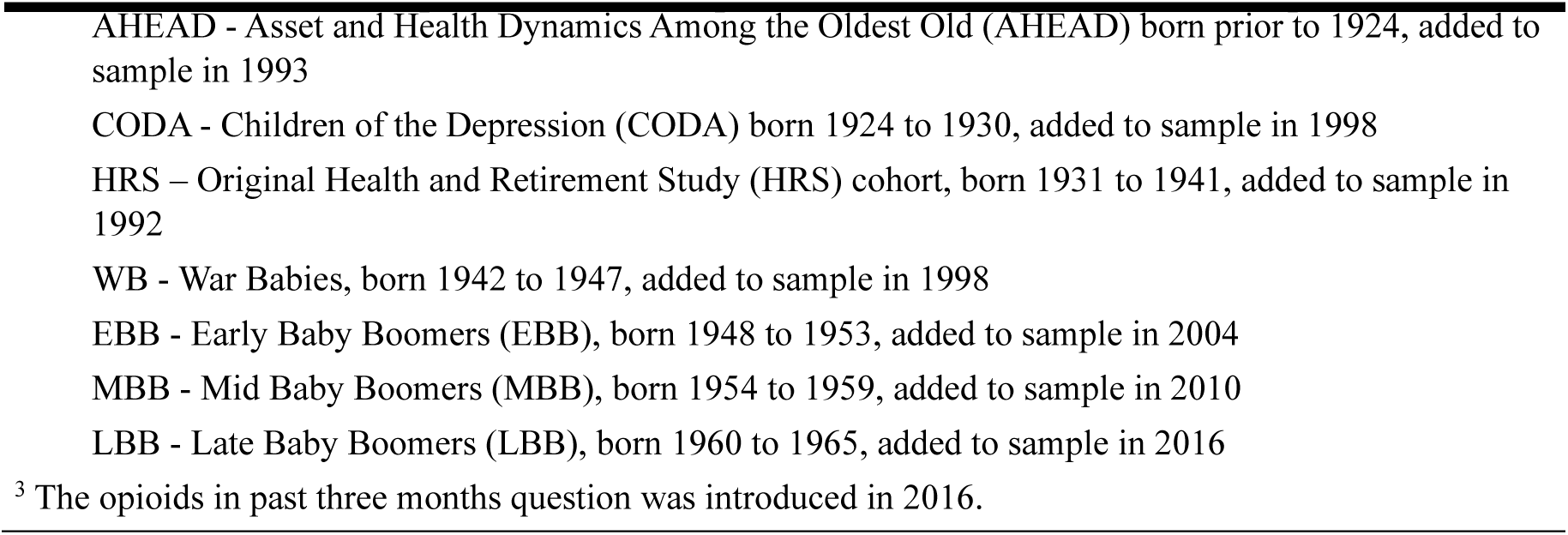
Demographics.

Pain is common in this population, with 36.8% of responses indicating frequent pain and 23.7% reporting pain that limits activities (Table 2). On average, respondents reported 2.6 functional limitations. In the surveys administered in 2016 and later, 13.0% reported opioid use in the past three months. No pre-implementation trends were detected for any of the outcomes (Figure 1).

**Figure 1:**
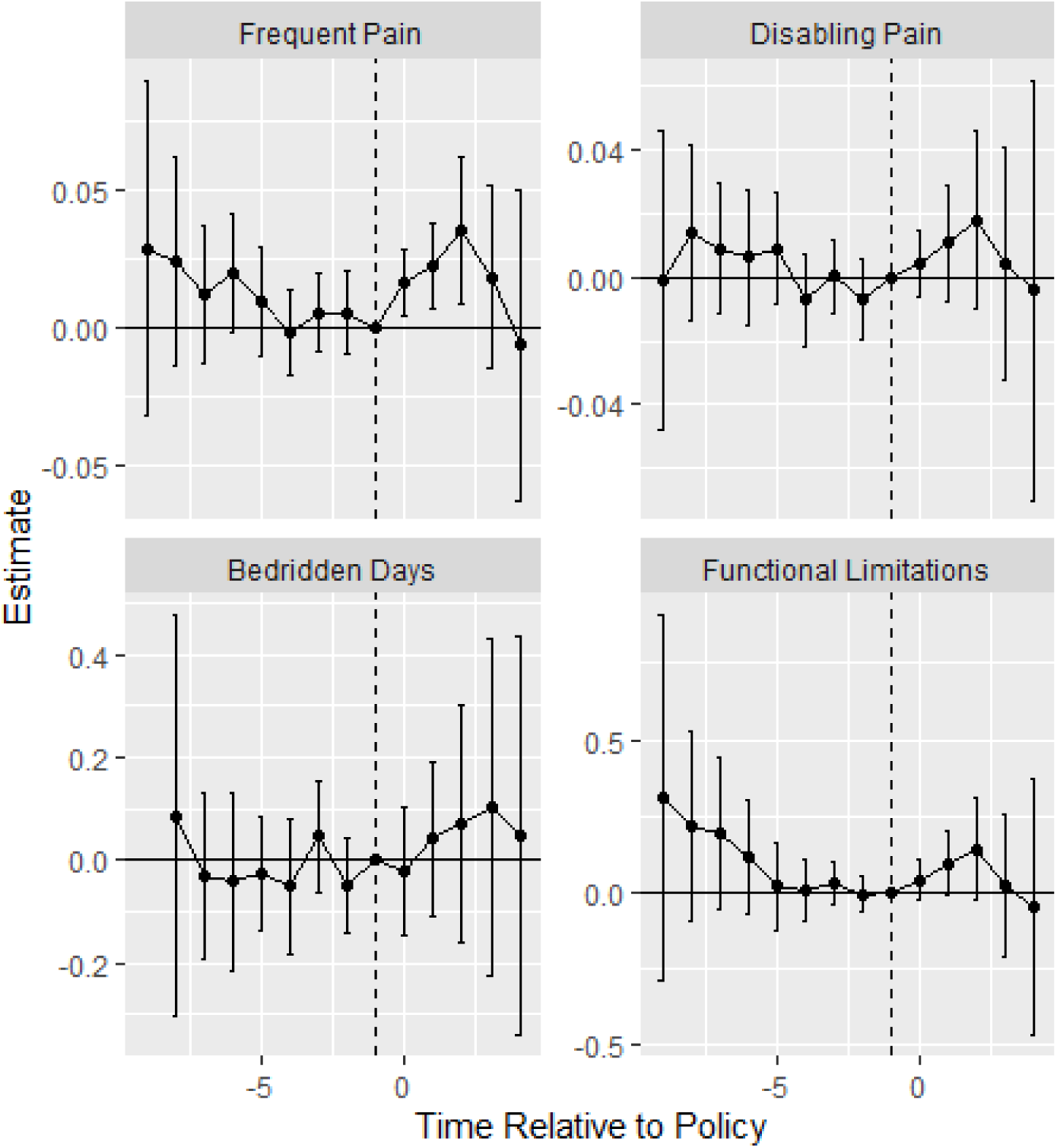
Effect of Must-Access PDMPs on Pain Measures.

**Table 2:**
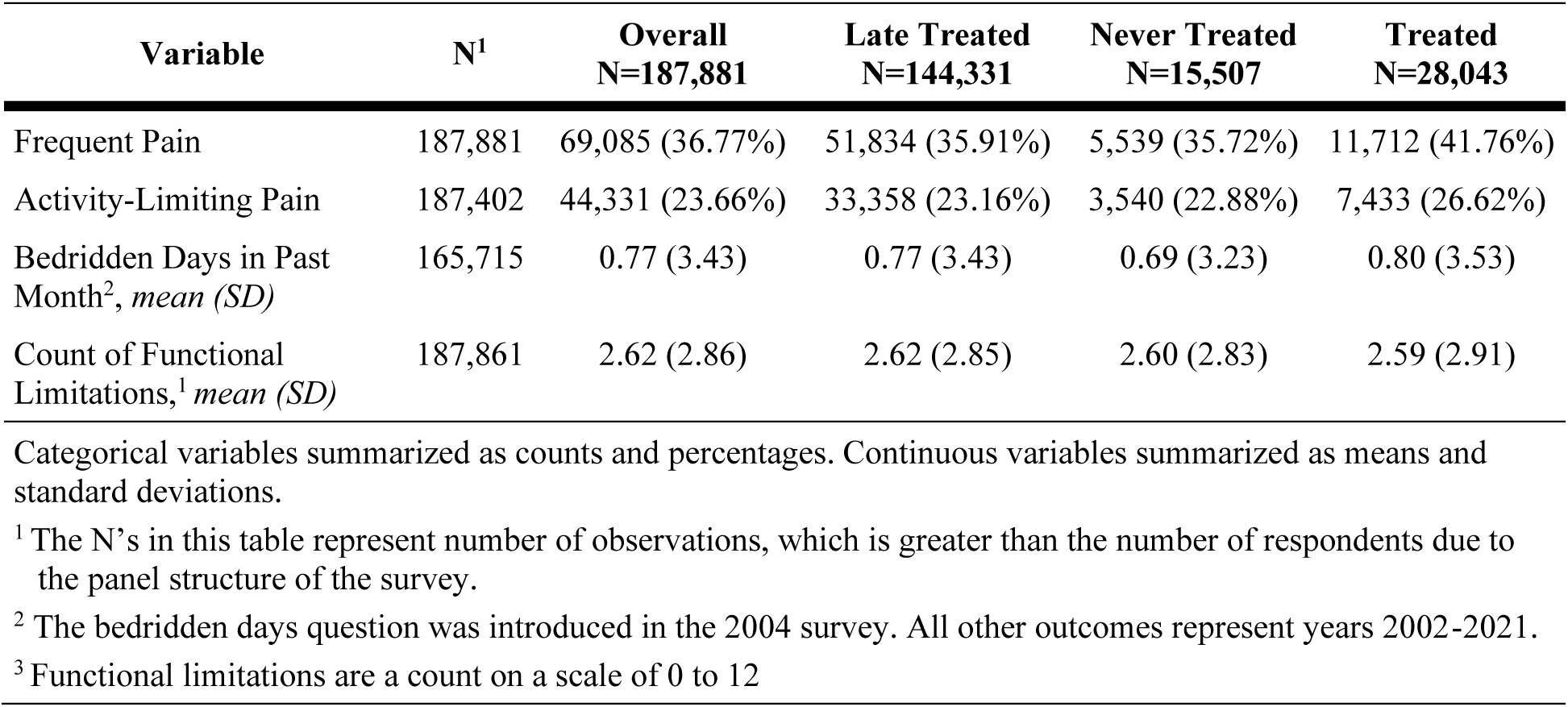
Unadjusted Outcome Summary.

For all outcomes, the average treatment effect aggregated across all post-policy periods was insignificant (Table 3). Underlying these insignificant aggregated effects was substantial heterogeneity across time periods (Figure 1). Must-access PDMPs were associated with an insignificant 1.7 (SE: 1.0, 95% CI: −0.3 - 3.7) percentage point increase in the percent of respondents reporting frequent pain. However, when disaggregated by time period, a significant increase in the percentage of respondents with frequent pain emerges in the policy implementation period and the two following periods. This increase peaks in the second post-policy period at 3.5 percentage points (SE: 1.0, 95% CI: 0.88 – 6.16). In subsequent time periods, the effect decreases in magnitude and becomes statistically indistinguishable from zero.

**Table 3:** Average Effect on the Treated.

| Statistics | Outcome |  |  |  |
| --- | --- | --- | --- | --- |
|  | Frequent Pain | Activity-Limiting Pain | Bedridden Days | Functional Limitations |
| Policy Effect (SD) | 0.017 (0.010) | 0.007 (0.011) | 0.048 (0.069) | 0.048 (0.059) |
| Number survey responses | 172,374 | 171,931 | 152,110 | 172,356 |

The results for the activity-limiting pain, bedridden days, and functional limitations models followed a similar pattern, but neither the aggregated nor the relative time effect estimates from the main models were statistically significant. Using the never-treated population as the comparison group instead of the late-treated population produced similar results (Supplemental Table 4).

## Discussion

Our results highlight the complexity of PDMP policy and pain treatment, along with the dynamic nature of policy in the opioid epidemic. We do not find significant overall effects on any of our outcomes. However, we find evidence for an increase in frequent pain in the first three periods following policy implementation. These effects appear to dissipate over time, although this finding should be interpreted with caution since a relatively small number of must-access PDMPs were implemented early enough to allow for more than three follow-up measurements using these data. Similar but statistically insignificant patterns are observed in the activity-limiting pain, functional limitations, and bedridden days outcomes.

Pain of any severity was reported by 36.8 percent of the sample and of activity-limiting pain was reported by 23.7 percent. This high prevalence aligns with prior research documenting upward trends in reported pain of all severity levels in the HRS population over time (Zimmer & Zajacova, 2020). The finding that must-access PDMPs were associated with an increase in frequent pain in the 4-6 years following policy implementation is likely driven by PDMPs leading to changes in the pain care environment, such as fewer pain relief options being offered by the clinician.

The effect estimates for the three outcomes that capture more severe pain and physical impairment were positive but not statistically significant. While the qualitative literature has described patients reporting increased disability and quality of life following opioid-restricting policies (Antoniou et al., 2019), such cases appear to be sufficiently rare among our studied population of older adults that they do not produce a clear signal in these data. Targeted research on chronic pain patients may provide more conclusive evidence on the population-specific effects of must-access PDMPs. As the 2022 update to the 2016 CDC Guideline for Prescribing Opioids for Chronic Pain acknowledged, tapering or discontinuing opioids in patients who have taken them long term can be associated with clinically significant risks, particularly if opioids are tapered rapidly or patients do not receive effective support (Dowell et al., 2022). In addition, patients may be reluctant to adopt a new treatment regime, believe it to be inferior to their opioid therapy, or have trouble adjusting to it. Overall, the transition away from opioids presents multiple opportunities for patients on long-term opioid therapy to experience increases in pain (Dowell et al., 2022).

Furthermore, our results are specific to older adults and may not translate to the general population, particularly given that some research has shown differential effects of PDMPs for non-elderly commercially insured patients and elderly Medicare patients (Bao et al., 2021).

From a methods perspective, our results reinforce the importance of using the newer, heterogeneity-robust difference in different methods to analyze the effects of PDMPs. We find that broadly summarizing an overall PDMP effect oversimplifies the story. Many of the seminal PDMP evaluations (Dowell, Zhang, et al., 2016; Horwitz et al., 2021) were published before heterogeneity-robust methods were standard. Multiple literature reviews have noted conflicting results present in the PDMP evaluation literature (Hoppe et al., 2022; Puac-Polanco et al., 2020; Tay et al., 2023), some of which may be explained by inconsistent PDMP definitions (Horwitz et al., 2021). Given our finding that the PDMP effect on pain frequency changed over time, the lack of heterogeneity-robust methods may be an additional contributor to the variety of conclusions present in the PDMP literature.

A limitation of this study is that the data did not allow measuring past opioid use. The HRS did not introduce an opioid use question until 2016, which was several years after the first wave of must-access PDMP policies went into effect. Although the HRS can be linked to Part D prescription data, less than 15 percent of HRS respondents have both Medicare Part D coverage and consented to the data linkage.

An additional limitation is that the opioid and pain policy environment was evolving during this time period. As mentioned above, the CDC prescribing guidelines for chronic pain were released in 2016 (Dowell, Haegerich, et al., 2016). The difference-in-difference structure of the analysis should eliminate a global effect of the post-guideline years. However, the guidelines may have interacted with existing PDMP policies in an unknown manner. Prescription limit laws also began to appear during this time. Laws that limit opioid prescription length to less than 30 days first began appearing in 2016 (Davis et al., 2019). We expect that the effect of these laws on our outcomes was minimal, given that the majority of these laws allow broad exceptions for clinician judgement and prior research has not shown an association between these laws and opioid distribution (Davis et al., 2020).

Marijuana laws comprised another area of active state-level policy changes. Medical marijuana laws have been shown to be associated with decreased pain, increased self-reported health, and increased labor market participation among older adults (Nicholas & Maclean, 2019). Three states implemented medical marijuana laws and must-access PDMPs within the same survey wave (i.e., two-year period). The effects of the must-access PDMPs may be suppressed by the effects of the medical marijuana laws in those states.

## Conclusion

This study is the first to evaluate the effect of must-access PDMPs on patient-centered measures of pain and physical impairment. Our analysis demonstrated initial increases in the percentage of older adults reporting frequent pain following policy implementation. All of the physical impairment outcomes were positive but insignificant, suggesting that increases in disabling pain following must-access PDMP implementation were not widespread in this population. These results highlight the importance of using heterogeneity-robust methods when assessing opioid policy effects.

## Supporting information

Supplemental Content

## Acknowledgements

The authors thank Sara Markowitz and Ian McCarthy for helpful feedback on early versions of the manuscript.

## Statements, Declarations, and Acknowledgements

### Ethical Approval

This study was deemed exempt by the Emory University Institutional Review Board on June 29, 2021 (IRB #STUDY00002935).

### Data Availability

The Health and Retirement (HRS) restricted access data, along with the must-access prescription drug monitoring program (PDMP) law data set created by the authors for this project, are housed in a secure data enclave hosted by the Michigan Center for the Demography of Aging (MiCDA). The data can be accessed by investigators with an approved Restricted Data Agreement with the University of Michigan. The PDMP law data set also is included in the appendix of the paper.

## Works Cited

Agree, E. M., & Wolf, D. A. (2017). Disability Measurement in the Health and Retirement Study. Forum Health Econ Policy, 21(1). 10.1515/fhep-2017-0029

Anson, P. (2017). Survey Finds CDC Opioid Guidelines Harming Patients. Pain News Network. Retrieved March 15 from https://www.painnewsnetwork.org/stories/2017/3/13/survey-finds-cdc-opioid-guidelines-harming-patients

Antoniou, T., Ala-Leppilampi, K., Shearer, D., Parsons, J. A., Tadrous, M., & Gomes, T. (2019). "Like being put on an ice floe and shoved away": A qualitative study of the impacts of opioid-related policy changes on people who take opioids. Int J Drug Policy, 66, 15–22. 10.1016/j.drugpo.2019.01.015

Bao, Y., Zhang, H., Wen, K., Johnson, P., Jeng, P. J., Witkin, L. R., Nicholson, S., Reid, M. C., & Schackman, B. R. (2021). Robust Prescription Monitoring Programs and Abrupt Discontinuation of Long-term Opioid Use. Am J Prev Med. 10.1016/j.amepre.2021.04.019

Benjamin, E. J., Virani, S. S., Callaway, C. W., Chamberlain, A. M., Chang, A. R., Cheng, S., Chiuve, S. E., Cushman, M., Delling, F. N., Deo, R., de Ferranti, S. D., Ferguson, J. F., Fornage, M., Gillespie, C., Isasi, C. R., Jiménez, M. C., Jordan, L. C., Judd, S. E., Lackland, D.,… Muntner, P. (2018). Heart Disease and Stroke Statistics-2018 Update: A Report From the American Heart Association. Circulation, 137(12), e67–e492. 10.1161/cir.0000000000000558

Coleman, C., Lennon, R. P., Garza, R. H., Veasley, C., Kuchera, J., Edwards, R., & Zgierska, A. E. (2023). Shifting quality chronic pain treatment measures from processes to outcomes. J Opioid Manag, 19(7), 83–94. 10.5055/jom.2023.0802

Colloca, L. (2019). The Placebo Effect in Pain Therapies. Annual Review of Pharmacology and Toxicology, 59(Volume 59, 2019), 191-211. 10.1146/annurev-pharmtox-010818-021542

Covinsky, K. E., Lindquist, K., Dunlop, D. D., & Yelin, E. (2009). Pain, functional limitations, and aging. J Am Geriatr Soc, 57(9), 1556–1561. 10.1111/j.1532-5415.2009.02388.x

Dahlhamer, J., Lucas, J., Zelaya, C., Nahin, R., Mackey, S., DeBar, L., Kerns, R., Von Korff, M., Porter, L., & Helmick, C. (2018). Prevalence of Chronic Pain and High-Impact Chronic Pain Among Adults — United States, 2016. Retrieved from https://www.cdc.gov/mmwr/volumes/67/wr/mm6736a2

Darnall, B. D., Juurlink, D., Kerns, R. D., Mackey, S., Van Dorsten, B., Humphreys, K., Gonzalez-Sotomayor, J. A., Furlan, A., Gordon, A. J., Gordon, D. B., Hoffman, D. E., Katz, J., Kertesz, S. G., Satel, S., Lawhern, R. A., Nicholson, K. M., Polomano, R. C., Williamson, O. D., McAnally, H.,… Lovejoy, T. (2019). International Stakeholder Community of Pain Experts and Leaders Call for an Urgent Action on Forced Opioid Tapering. Pain Medicine, 20(3), 429–433. 10.1093/pm/pny228

Davis, C. S., Lieberman, A. J., Hernandez-Delgado, H., & Suba, C. (2019). Laws limiting the prescribing or dispensing of opioids for acute pain in the United States: A national systematic legal review. Drug and Alcohol Dependence, 194, 166–172. 10.1016/j.drugalcdep.2018.09.022

Davis, C. S., Piper, B. J., Gertner, A. K., & Rotter, J. S. (2020). Opioid Prescribing Laws Are Not Associated with Short-term Declines in Prescription Opioid Distribution. Pain Medicine, 21(3), 532–537. 10.1093/pm/pnz159

Dobscha, S. K., Corson, K., Flores, J. A., Tansill, E. C., & Gerrity, M. S. (2008). Veterans affairs primary care clinicians’ attitudes toward chronic pain and correlates of opioid prescribing rates. Pain Med, 9(5), 564–571. 10.1111/j.1526-4637.2007.00330.x

Dowell, D., Haegerich, T. M., & Chou, R. (2016). CDC guideline for prescribing opioids for chronic pain—united states, 2016. JAMA, 315(15), 1624–1645. 10.1001/jama.2016.1464

Dowell, D., Ragan, K., Jones, C., Baldwin, G., & Chou, R. (2022). CDC Clinical Practice Guideline for Prescribing Opioids for Pain — United States, 2022. Retrieved from 10.15585/mmwr.rr7103a1

Dowell, D., Zhang, K., Noonan, R. K., & Hockenberry, J. M. (2016). Mandatory Provider Review And Pain Clinic Laws Reduce The Amounts Of Opioids Prescribed And Overdose Death Rates. Health Aff (Millwood), 35(10), 1876–1883. 10.1377/hlthaff.2016.0448

Fenton, J. J., Agnoli, A. L., Xing, G., Hang, L., Altan, A. E., Tancredi, D. J., Jerant, A., & Magnan, E. (2019). Trends and Rapidity of Dose Tapering Among Patients Prescribed Long-term Opioid Therapy, 2008-2017. JAMA Network Open, 2(11), e1916271–e1916271. 10.1001/jamanetworkopen.2019.16271

Finley, E. P., Garcia, A., Rosen, K., McGeary, D., Pugh, M. J., & Potter, J. S. (2017). Evaluating the impact of prescription drug monitoring program implementation: a scoping review. BMC Health Services Research, 17, 420. 10.1186/s12913-017-2354-5

Florence, C., Luo, F., & Rice, K. (2021). The economic burden of opioid use disorder and fatal opioid overdose in the United States, 2017. Drug Alcohol Depend, 218, 108350. 10.1016/j.drugalcdep.2020.108350

Fornili, K. (2018). The Opioid Crisis, Suicides, and Related Conditions: Multiple Clustered Syndemics, not Singular Epidemics. J Addict Nurs, 29(3), 214–220. 10.1097/jan.0000000000000240

Gaskin, D. J., & Richard, P. (2012). The Economic Costs of Pain in the United States. The Journal of Pain, 13(8), 715–724. 10.1016/j.jpain.2012.03.009

Gleason, R. M., K. L. Kirsh, S. D. Passik, and J. F. Chambers. (2014). Current Access to Opioids: Survey of Chronic Pain Patients. Practical Pain Management.

Goodin, A. J. (2018). We Cannot Treat the Dead. Am J Public Health, 108(10), 1286–1288. 10.2105/ajph.2018.304658

Gross, J., & Gordon, D. B. (2019). The Strengths and Weaknesses of Current US Policy to Address Pain. Am J Public Health, 109(1), 66–72. 10.2105/ajph.2018.304746

Health and Retirement Study. (2017). Aging in the 21st Century: Challenges and Opportunities for Americans. https://hrsonline.isr.umich.edu/sitedocs/databook/inc/pdf/HRS-Aging-in-the-21St-Century.pdf

Health and Retirement Study. (2025). Produced and distributed by the University of Michigan with funding from the National Institute on Aging (grant number NIA U01AG009740).

Heeringa, S., & Connor, J. (1995). Technical Description of the Health and Retirement Survey Sample Design. https://hrs.isr.umich.edu/publications/biblio/5310

Holmgren, A. J., Botelho, A., & Brandt, A. M. (2020). A History of Prescription Drug Monitoring Programs in the United States: Political Appeal and Public Health Efficacy. Am J Public Health, 110(8), 1191–1197. 10.2105/ajph.2020.305696

Hoppe, D., Karimi, L., & Khalil, H. (2022). Mapping the research addressing prescription drug monitoring programs: A scoping review. Drug Alcohol Rev, 41(4), 803–817. 10.1111/dar.13431

Horwitz, J. R., Davis, C., McClelland, L., Fordon, R., & Meara, E. (2021). The importance of data source in prescription drug monitoring program research. Health Services Research, 56(2), 268–274. 10.1111/1475-6773.13548

Human Rights Watch. (2018). Not Allowed to Be Compassionate: Chronic Pain, the Overdose Crisis, and Unintended Harms in the US. https://www.hrw.org/report/2018/12/18/not-allowed-be-compassionate/chronic-pain-overdose-crisis-and-unintended-harms-us

Kilby, A. E. (2015). Opioids of the masses: Welfare tradeoffs in the regulation of narcotic pain medications. Massachusetts Institute of Technology. http://economics.mit.edu/files/11150

Komro, K. A., O’Mara, R. J., & Wagenaar, A. C. (2013). Public health law research : Theory and methods. In A. C. Wagenaar & S. C. Burris (Eds.). John Wiley & Sons, Incorporated.

National Academy of Sciences. (2017). Pain Management and the Opioid Epidemic: Balancing Societal and Individual Benefits and Risks of Prescription Opioid Use. Washing, DC: The National Academies Press

Nicholas, L. H., & Maclean, J. C. (2019). The Effect of Medical Marijuana Laws on the Health and Labor Supply of Older Adults: Evidence from the Health and Retirement Study. Journal of Policy Analysis and Management, 38(2), 455–480. 10.1002/pam.22122

Peachman, R. R. (2023). Will the New CDC Opioid Prescribing Guidelines Help Correct the Course in Pain Care? JAMA, 329(2), 111–113. 10.1001/jama.2022.22284

Picco, L., Lam, T., Haines, S., & Nielsen, S. (2021). How prescription drug monitoring programs influence clinical decision-making: A mixed methods systematic review and meta-analysis. Drug and Alcohol Dependence, 228, 109090. 10.1016/j.drugalcdep.2021.109090

Prescription Drug Monitoring Program Training and Technical Assistance Center. (2024). Mandatory PDMP Use. Retrieved March 29 from https://www.pdmpassist.org/Content/Documents/pdf/Mandatory_Enrollment_Conditions.pdf

Puac-Polanco, V., Chihuri, S., Fink, D. S., Cerdá, M., Keyes, K. M., & Li, G. (2020). Prescription Drug Monitoring Programs and Prescription Opioid-Related Outcomes in the United States. Epidemiol Rev. 10.1093/epirev/mxaa002

RAND-USC Schaeffer Opioid Policy Tools and Information Center. (2021a). OPTIC-Vetted Marijuana Policy Data. https://www.rand.org/health-care/centers/optic/resources/datasets.html

RAND-USC Schaeffer Opioid Policy Tools and Information Center. (2021b). OPTIC-Vetted PDMP Policy Data. Retrieved 10/20/21 from https://www.rand.org/health-care/centers/optic/resources/datasets.html

Sehgal, N., Colson, J., & Smith, H. S. (2013). Chronic pain treatment with opioid analgesics: benefits versus harms of long-term therapy. Expert Rev Neurother, 13(11), 1201–1220. 10.1586/14737175.2013.846517

Sherman, K. J., Walker, R. L., Saunders, K., Shortreed, S. M., Parchman, M., Hansen, R. N., Thakral, M., Ludman, E. J., Dublin, S., & Von Korff, M. (2018). Doctor-Patient Trust Among Chronic Pain Patients on Chronic Opioid Therapy after Opioid Risk Reduction Initiatives: A Survey. J Am Board Fam Med, 31(4), 578–587. 10.3122/jabfm.2018.04.180021

Sonnega, A., Faul, J. D., Ofstedal, M. B., Langa, K. M., Phillips, J. W., & Weir, D. R. (2014). Cohort Profile: the Health and Retirement Study (HRS). Int J Epidemiol, 43(2), 576–585. 10.1093/ije/dyu067

Substance Abuse and Mental Health Services Administration. (2024). Key Substance Use and Mental Health Indicators in the United States: Results from the 2023 National Survey on Drug Use and Health. (HHS Publication No. PEP24-07-021, NSDUH Series H-59). Retrieved from https://www.samhsa.gov/data/report/2023-nsduh-annual-national-report

Szalavitz, M. (2022, Mar 22, 2022). What the Opioid Crisis Took From People in Pain. The New York Times. https://www.nytimes.com/2022/03/07/opinion/opioid-crisis-pain-victims.html

Tay, E., Makeham, M., Laba, T. L., & Baysari, M. (2023). Prescription drug monitoring programs evaluation: A systematic review of reviews. Drug Alcohol Depend, 247, 109887. 10.1016/j.drugalcdep.2023.109887

Tompkins, D. A., Hobelmann, J. G., & Compton, P. (2017). Providing chronic pain management in the “Fifth Vital Sign” Era: Historical and treatment perspectives on a modern-day medical dilemma. Drug and Alcohol Dependence, 173, S11–S21. 10.1016/j.drugalcdep.2016.12.002

Upshur, C. C., Bacigalupe, G., & Luckmann, R. (2010). "They don’t want anything to do with you": patient views of primary care management of chronic pain. Pain Med, 11(12), 1791–1798. 10.1111/j.1526-4637.2010.00960.x

Wetzel, M., von Esenwein, S., Yarbrough, C. R., & Hockenberry, J. M. (2021). Association of Prescription Drug Monitoring Program Laws with Bedridden and Missed Work Days. Health Serv Res, 1–7.

White, A. P., Arnold, P. M., Norvell, D. C., Ecker, E., & Fehlings, M. G. (2011). Pharmacologic management of chronic low back pain: synthesis of the evidence. Spine (Phila Pa 1976), 36(21 Suppl), S131–143. 10.1097/BRS.0b013e31822f178f

Zimmer, Z., & Zajacova, A. (2020). Persistent, Consistent, and Extensive: The Trend of Increasing Pain Prevalence in Older Americans. J Gerontol B Psychol Sci Soc Sci, 75(2), 436–447. 10.1093/geronb/gbx162

