## Supplemental Content for "Do Must-Access Prescription Drug Monitoring Programs (PDMPs) Affect Pain and Physical Impairment Outcomes in Older Adults?"

**Supplemental Table 1: Must-Access PDMP Policy Dates**

| State | Policy Date |
|---|---|
| Alabama | 3/1/2017 |
| Alaska | 7/17/2017 |
| Arizona | 10/1/2017 |
| Arkansas | 8/1/2017 |
| California | 10/2/2018 |
| Colorado | 9/7/2021 |
| Connecticut | 10/1/2015 |
| Delaware | N/A |
| District of Columbia | 1/14/2021 |
| Florida | 12/19/2018 |
| Georgia | 7/1/2018 |
| Hawaii | 7/1/2018 |
| Idaho | 10/1/2020 |
| Illinois | 1/1/2018 |
| Indiana | 1/1/2021 |
| Iowa | 11/27/2019 |
| Kansas | N/A |
| Kentucky | 7/20/2012 |
| Louisiana | 2/1/2018 |
| Maine | 1/1/2017 |
| Maryland | 7/1/2018 |
| Massachusetts | 12/5/2014 |
| Michigan | 6/1/2018 |
| Minnesota | N/A |
| Mississippi | N/A |
| Missouri | N/A |
| Montana | 7/1/2021 |
| Nebraska | N/A |
| Nevada | 5/5/2015 |
| New Hampshire | 1/1/2016 |
| New Jersey | 9/1/2015 |
| New Mexico | 1/1/2017 |
| New York | 8/27/2013 |
| North Carolina | 7/1/2021 |
| North Dakota | 1/1/2018 |
| Ohio | 4/1/2015 |
| Oklahoma | 11/15/2015 |
| Oregon | 9/15/2021 |
| Pennsylvania | 8/18/2016 |
| Rhode Island | 3/1/2015 |
| South Carolina | 5/19/2017 |
| South Dakota | N/A |

| State | Policy Date |
| --- | --- |
| Tennessee | 4/1/2013 |
| Texas | 3/1/2020 |
| Utah | 5/8/2018 |
| Vermont | 11/15/2013 |
| Virginia | 7/1/2015 |
| Washington | 10/1/2021 |
| West Virginia | 5/1/2013 |
| Wisconsin | 4/1/2017 |
| Wyoming | 3/12/2020 |

**Supplemental Table 2: Functional Limitations Measure:**

| Functional Limitations |
| --- |
| Running/jogging about 1 mile |
| Walking several blocks |
| Walking 1 block |
| Sitting for about 2 hours |
| Getting up from a chair |
| Climbing several flights of stairs |
| Climbing one flight of stairs |
| Lifting or carrying over 10 lbs. |
| Stooping, crouching or kneeling |
| Picking a dime up |
| Reaching or extending arms |
| Pulling or pushing large objects |

**Supplemental Table 3: Survey Questions**

| Role | Construct | Full Question | Variable | Cleaning/Edits |
|---|---|---|---|---|
| Independent variable | State | State of residence | STFIPS | N/A |
| Outcome | Frequent pain | Are you often troubled with pain? | C104 | N/A |
| Outcome | Activity-limiting pain | Does the pain make it difficult for you to do your usual activities such as household chores or work? | C106 | Outcome positive if C106=Yes, negative if C106=No or C104=No |
| Outcome | Bedridden days | Aside from any hospital or nursing home stays, about how many days did you stay in bed more than half the day because of illness or injury during the last month? | C229 | Set to missing if outside range 0-31 |
| Outcome | Functional limitations | Functional Limitations and Helpers-Section_G: CHKPNT: COUNT OF G001 THROUGH G012 | G013 | N/A |

**Supplemental Table 4: Never Treated as Comparison Group**

| Statistic | Frequent Pain | Activity-Limiting Pain | Bedridden Days | Functional Limitations |
|---|---|---|---|---|
| Effect | 0.011 | 0.002 | 0.061 | 0.040 |
| SE | 0.012 | 0.012 | 0.070 | 0.079 |
| Number survey responses | 43,550 | 43,392 | 41,088 | 43,548 |
